# Deep Learning-based Framework for Rapid, Precise and Fully Automated Localization of Cochlea Landmarks in 3D CT Images

**DOI:** 10.64898/2026.11.06.26366894

**Authors:** Yifan Wang, Andrej Kral, Thomas Lenarz, Samuel John

## Abstract

Manual localization of anatomical landmarks within human cochlea structures in 3D computed tomography (CT) images is labor-intensive. However, despite progress in various studies on automatic localization methods, it remains challenging to jointly achieve rapid, precise, and fully automated landmark localization. Therefore, we propose a deep learning-based localization framework, called MEDICO-YOLO, which bridges speed and precision. It can automatically and simultaneously localize 6 human cochlea landmarks (apex, basal point, round window center of both left and right cochleae) in the full-resolution 3D CT images. The main contributions are: 1) MEDICO-YOLO learns from few manually annotated 3D CT images; 2) Within the framework, a 2D CNN-based network was developed to specifically localize cochlea landmarks in 3D single-scale medical CT images; 3) A novel loss function was designed; 4) The localization model achieved human expert-level precision while ensuring rapid inference. This study employed anonymized 3D pre-operative CT images (0.3 mm, covering the full head or temporal bone region). The evaluation involved comparing four models that were trained using 4 different training set sizes (n=4, 8, 12, 16). Each of them was evaluated on a distinct test set (n=16). The results show that the MEDICO-YOLO framework achieved strong localization performance by learning from few annotated 3D CT images. Trained on only 8 CT volumes, the model already attained a mean localization error of 1.30 mm (approximately 4 voxels). When trained on 16 CT volumes, the model achieved a mean error of 0.71 mm and a successful detection rate (SDR@3.0 mm) of 97.92%. Furthermore, on an Apple M1 laptop, the model achieved a real-time speed (32 ms) for inferencing each 2D slice (608 × 608) and an average latency of 25.43 s for inferencing the full 3D CT image (608 × 608 × 200, including post-processing). The complete methodology for the MEDICO-YOLO framework is presented in this article.

## INTRODUCTION

Understanding the precise spatial information of patient-specific anatomical structures in medical imaging is critical for individualized surgical planning and other medical applications. Relevant to the temporal bone region, different studies have focused on the segmentation of anatomical structures (1,2). However, in cochlear implant surgery, segmenting the shape of a structure alone is often insufficient, localizing clinically relevant landmarks (e.g., cochlea apex, basal point and round window center) can also be crucial for performing minimally invasive surgical procedures (3). Whereas manually localizing cochlea landmarks remains challenging and labor-intensive due to the cochlea’s minute size and significant inter-individual variation (4,5).

Earlier studies employed atlas-based approaches for automatic localization or segmentation of anatomical structures within the temporal bone region (6–8). Such approaches employ predefined anatomical atlas constructed from annotated dataset as templates, aligning them with new patient scans via registration techniques, demonstrating precise localization performance. However, it is often sensitive to anatomical variability and can become computationally expensive when integrating multi-atlas strategy or deformable registration technique.

Deep learning-based convolutional neural networks (CNNs) have been increasingly employed in various studies to perform automatic landmark detection in medical images (9,10). More relevant to our domain, several studies have proposed CNN-based approaches for localizing anatomical structures within the temporal bone region. Margeta et al. (11) developed the web-based platform Nautilus, employing a 3D U-Net architecture for cochlea landmark detection and segmentation in pre- and post-operative images. For detection in pre-operative images, the 3D U-Net outputs a heatmap for each landmark, 3 heatmaps in total. Subsequently, the coordinates of landmarks are extracted by identifying peak responses on the predicted Gaussian probability maps. The network trained with 279 pre- and post-operative scans, achieved an average localization error of approximately 2 voxels on pre-operative CT images. However, this method requires a manual initialization via a user-defined 10 × 10 × 10 mm region of interest (ROI). Wang et al. (12) demonstrated an approach to train the algorithm using only a single annotated CT scan. They first employed a shallow 2D U-Net to segment the structure of interest (SOI) mask, enabling an initial alignment between the target image and reference image. Subsequently, they integrated 3D registration to refine the coordinates of landmarks. However, the accuracy of this method can be sensitive to the quality of the single annotated reference image, and the registration-based optimization can become complex when large anatomical variation presents. Additionally, Stebani et al. (13) proposed a dual-headed 3D U-Net trained on 38 CT scans to achieve both temporal bone segmentation and landmark detection. In the detection component, they integrated a heatmap regression output layer into the network’s head module and implemented its corresponding regression loss. This approach rapidly localizes the helicotrema, oval window and round window within a 128 × 128 × 128 voxel sub-volume, achieving an average localization error of 5.2 voxel units. Furthermore, Wang et al. (14) proposed a few-shot learning approach for automatic localization of several cochlea landmarks (apex, basal point and round window center), combining an optimized data pre-processing operation with a tailored multi-class classification 2D CNN model. A sliding window strategy was then employed at the localization process. Using only 5 annotated CT scans as training data, their algorithm achieved an average localization error of approximately 3 voxels on the input 3D CT volumes. However, due to its two-stage design, a manual ROI initialization is required to accelerate the inference.

These works underscore that the landmarks within the temporal bone area can be localized automatically and precisely on 3D CT images. However, due to multi-stage strategies or the characteristic of the U-Net architecture in localization (which often outputs one heatmap with full input image size per landmark), some approaches necessitate a manually defined ROI to accelerate the detection process. Achieving both rapid and precise automatic landmark localization on CT images without pre-cropping ROI, still remains a challenge.

Two-stage object detection algorithms such as R-CNN, Fast R-CNN, Faster R-CNN (15–17) first require filtering ROIs through a region proposal mechanism, then followed by the object classification and bounding box regression on the selected ROIs. Although these methods can achieve high detection precision, such two-stage detection design still slows down the inference speed. On the other hand, single-stage detector like center-based approach CenterNet (18) can predict directly the object’s center with heatmap-based output. However, each class necessitates a separate heatmap to detect its center, increasing computational cost during inference.

Another single-stage detection algorithm, You Only Look Once (YOLO) (19), has gained more attention owing to its real-time inference capability, global context and end-to-end processing pipeline. YOLO employs a single grid-based CNN model to process the entire input image in a single forward pass. By treating object detection as regression rather than classification, it simultaneously predicts the class, confidence scores and bounding box coordinates of multiple presented objects at output, thereby reducing inference duration while retaining high detection accuracy.

Unlike conventional YOLO, which primarily detects multi-scale objects on 2D natural images, our clinical task focuses on localizing anatomical landmarks within the human cochlea structure: the apex (helicotrema), the basal point (center of the cochlea basal turn), and the round window center. The dataset in use is 3D medical CT images covering the entire head or temporal bone region, with consistent isotropic image resolution.

Therefore, we propose a deep learning-based, automated landmark localization framework, named MEDICO-YOLO (MEDical Image Centers Only YOLO), which is single-scale, 3D medical CT image-focused. The methodology of this framework encompasses 2D CNN-based network architecture, loss function, data generation, training configuration, and the 3D localization inference pipeline. It supports few-volume learning and localizes multiple cochlea landmarks simultaneously and fully automatically, attains both high precision and rapid inference. Furthermore, it requires no manual initialization thanks to its single-stage design.

To the best of our knowledge, this is the first automated cochlea landmark localization framework that achieves collectively all of the aforementioned features. Thus, the main contributions include:

1. To enable few-volume learning while solving the overfitting issue, a custom data generator was developed. From a limited set of manually labeled 3D CT volumes, the data generator could generate a large and adjustable amount of 2D images and their corresponding ground truth label matrix for training and validation data. To enhance 3D spatial context on the generated 2D images, 3D augmentation was also integrated.
2. A 2D CNN-based network architecture was developed. Since the target cochlea structure appeared at a consistent scale on the studied dataset (isotropic image spacing of 0.3 mm), the neck module was omitted, and the backbone and head modules were adapted accordingly. Thus, the network outputs single-scale rather than multi-scale feature maps, thereby reducing the network size.
3. A novel loss function was designed from scratch, this included the implementation of customized individual loss components and an overall loss calculation mechanism that considers different cases. This loss function determined the model’s learning direction, contributing to its high precision. It also led to a correlation between the confidence threshold and precision, where the model localizes more precisely with a higher confidence threshold.
4. A localization inference pipeline integrated DBSCAN clustering (20) was developed to directly process the input full-resolution 3D CT scan. Without any manual initialization, it simultaneously outputs the 3D coordinates of all 6 target landmarks on either a full-head or temporal bone 3D CT scan.
5. The overall MEDICO-YOLO framework was evaluated quantitatively and qualitatively on a distinct test set (n=16), including inference speed, localization precision, qualitative visualization, comparison with human experts in localization precision, and overall comparison with other related works. The results demonstrate that this framework achieved human expert-level precision while maintaining rapid inference speed.

## MATERIAL AND METHODS

### Data Description

The dataset employed in this study (TABLE 1) consists of 36 anonymized preoperative 3D cone beam computed tomography (CBCT) scans, including 17 scans covering the human head and 19 scans covering the temporal bone region. The image shape for full head CT scan is 812 × 812 × 460, for temporal bone CT scan is 536 × 536 × 198. The dimension represents (X, Y, Z) in the axial view, where X and Y denote axial dimension, and Z denotes the number of axial slices. These scans were acquired from 36 patients (21 females and 15 males, all adults), using 2 different scanners, between 2010 to 2023. Each scan includes both left and right cochleae, with 0.3 mm isotropic image spacing and no gantry tilt. 21 of them were scanned at the ENT clinic or Neuroradiology department in Medizinische Hochschule Hannover with the institutional review board’s generic ethics decision No. 1897-2013 from 2013-07-09 regarding publications with patient data. Other 15 scans were acquired from full-body donors (ethics vote No. A 2016-0083) with University of Rostock. The scans with strong motion artifacts were excluded, while scans showing strong metal artifact on the side of the head (due to the attachment of a mini-stereotactic metal frame) were included. Ground truth annotations were manually performed by an expert using 3D Slicer (21), and divided into training, validation and testing sets, with each group containing distinct CT scans. To assess few-volume learning performance, the model was trained on 4 distinct training sets containing 4, 8, 12 and 16 manually labeled CT volumes respectively. For testing, a separate test set comprising 16 different CT volumes was employed.

**TABLE 1.** Distribution characteristics of datasets for training, validation and testing. TB: Temporal Bone; XT: Xoran Technologies; xCAT: xCAT® ENT.

| **Characteristic** | **Training Set (N=16)** | **Validation Set (N=4)** | **Test Set (N=16)** | **Total (N=36)** |
|---|---|---|---|---|
| Image Shape | 9 Full head, 7 TB | 2 Full head, 2 TB | 6 Full head, 10 TB | 17 Full head, 19 TB |
| Gender | 12 Female, 4 Male | 3 Female, 1 Male | 6 Female, 10 Male | 21 Female, 15 Male |
| CT Type | CBCT | CBCT | CBCT | CBCT |
| Manufacturer | XT | XT | XT | XT |
| Scanners | MiniCAT, xCAT | MiniCAT, xCAT | MiniCAT, xCAT | MiniCAT, xCAT |
| Resolution | 0.3 mm | 0.3 mm | 0.3 mm | 0.3 mm |

### Network Architecture

Since clinicians or radiologists commonly analyze 3D CT scans with 2D multiplanar views, we accordingly redefined the 3D CT cochlea landmark localization task as 2D slice localization in the axial direction. Therefore, a 2D CNN-based network architecture was developed. Compared to 3D convolutions, this 2D convolution-based adjustment reduces the computational burden by eliminating convolution operations along the depth dimension. The backbone module of the network was inspired by the CSPDarknet53 (22), employing Mish (23) as activation function. Beyond the backbone module, several architectural modifications tailored to our task were implemented: (1) The input image was adjusted to a single-channel image with the shape of 1 × 608 × 608. (2) The conventional neck module was removed due to the consistent anatomical size of the cochlea structure and fixed isotropic voxel spacing of 0.3 mm across the studied dataset, which eliminated the need for multi-scale feature fusion. (3) The head module was adjusted accordingly to the modification of the neck module, which outputs a single-scale feature map with an anchor-free design (24,25), and (4) the shape of the output tensor was tailored to 13 × 13 × 2 × 9.

As FIGURE 1 illustrates, the input image is downsampled across 5 successive CSP stages. Each stage integrates CSPNet (26), which divides the input feature map into two branches: one branch processes through a series of residual blocks (27), while the other bypasses them, the two branches are then merged via channel concatenation. This design reduces the computation while keeping both spatial and semantic information. Following the backbone module, the feature map is directly connected to the head module, which consists of five 3 × 3 convolution layer and one 1 × 1 convolution layer, Leaky ReLU is employed as activation function. The output tensor has a shape of 13 × 13 × 2 × 9, corresponding to 13 × 13 grid cells that divide the input image, each cell contains two bounding boxes, where each box consists of 9 parameters: x, y, w, h, confidence, and a class label represented by one-hot encoding (4 classes in total). The parameters (x, y, w, h) are defined relative to each grid cell.

**FIGURE 1.**
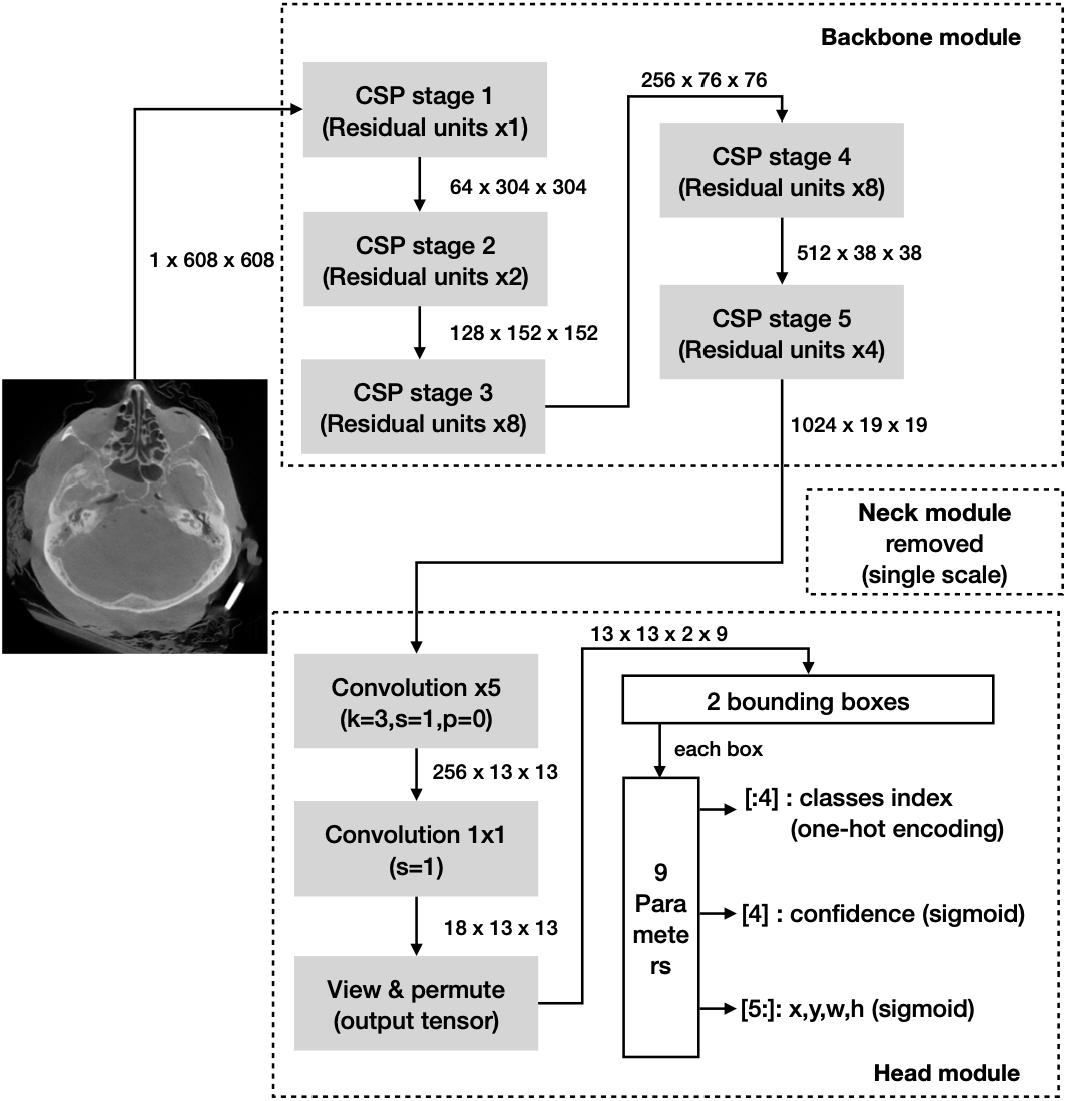
2D CNN network architecture in MEDICO-YOLO. Input image shape 1 × 608 × 608. The backbone module follows cspdarknet53. The neck module has been removed owing to the single-scale design. The head module has been modified accordingly, outputting a single scaled tensor of shape 13 × 13 × 2 × 9.

As anatomically, the human cochlea structure has a dimension of approximately 10 mm × 8 mm × 4 mm, which corresponds to 33 × 27 × 13 voxels in 0.3 mm resolution CT scan (4). Thus, to ensure the output feature map contains a sufficient receptive field, and each cell approximately covers the dimension of the cochlea structure, the size of input image was determined to 608 × 608, and the grid size to 13 × 13. In this single-scale design, such suitable spatial configuration is particularly important since all spatial information is presented on a single scale feature map.

### Loss Function

Beyond the network architecture, the definition of the loss function was equally crucial, as the network defines the feature extraction capacity while the loss function determines the learning objective. Thus, to localize cochlea landmarks, a novel loss function was designed. Unlike traditional YOLO family, we redesigned the loss function to directly predict the landmarks (x, y) coordinates rather than the bounding box, the (w, h) parameters were then omitted and thus eliminated the need of traditional loss metrics like intersection over union (IoU). Each cell was designed to predict 2 landmarks, which is consistent with the cochlea anatomy presented in the studied dataset. Each grid cell on the 2D axial CT slice can contain at most 2 cochlea landmarks (apex, basal point).

In the loss function, the prediction with the minimum matching cost (1) comparing to the ground truth, is responsible for localizing the corresponding landmark.

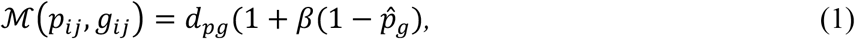

where *i* and *j* represent the index of the cells, *ℳ p*_*ij*_, *g*_*ij*_& was defined as the matching cost, encompassing both localization and classification metrics. *P*_*ij*_ denotes as predicted tensor and *g*_*ij*_ denotes as ground truth label, *d*_*pg*_ denotes the mean square error (MSE) between them. 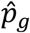 was defined as the softmax of the predicted class probability for the ground truth class *g*. Hyperparameter *β* was set to 0.17.

As shown in FIGURE 2, each input image corresponds to a 2D axial slice from a 3D CT image, which is divided by 13 × 13 cells. The predicted outputs for each cell were compared against the ground truth, the corresponding error and total loss were then calculated based on the designed loss calculation mechanism.

**FIGURE 2.**
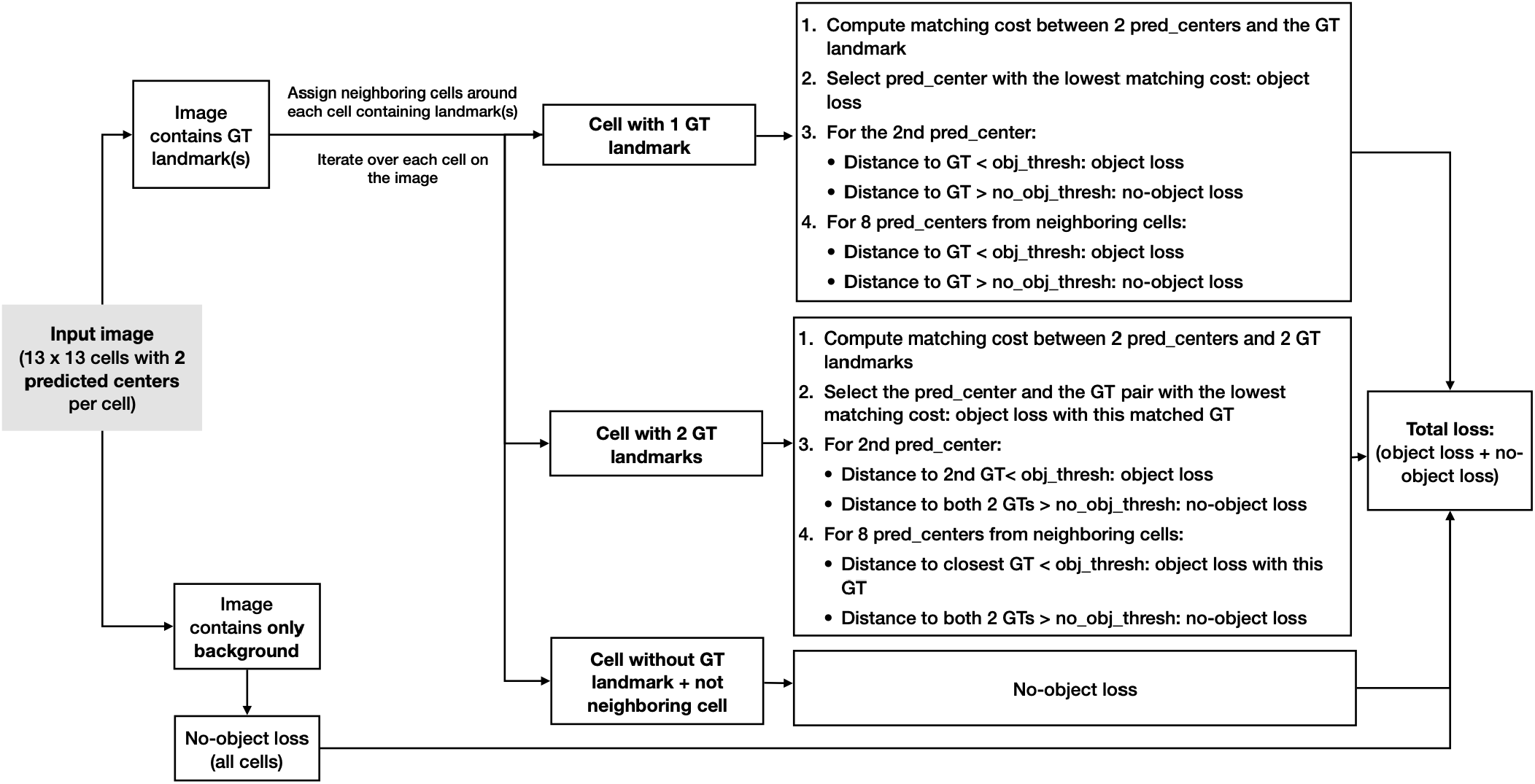
MEDICO-YOLO loss function diagram. Pred_center represents predicted center. GT represents ground truth landmark. The thresholds obj_thresh and no_obj_thresh define the distance criteria for matching the predicted and ground truth landmarks based on their Euclidean distance, obj_thresh was set to 0.16 and no_obj_thresh was set to 0.26.

For an image contains only background, no-object loss (2) was applied to all the predicted outputs,

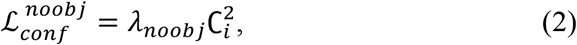

where 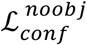 denotes the confidence loss component for the background cells, *λ*_*noobj*_ = 0.1 and *C*_*i*_ represents the predicted confidence score, and the target confidence for background cells was set to 0.

Since landmarks can locate close to cell boundaries, for an image containing ground truth landmarks, each cell containing a ground truth landmark was further assigned together with its 4 adjacent neighboring cells: (*i* + 1, *j*), (*i* − 1, *j*), (*i, j* + 1) and (*i, j* − 1). The predicted landmarks within the ground truth cell and its neighboring cells were all assigned to be responsible for the presented landmarks in the ground truth cell.

Based on the defined matching cost and Euclidean distance in pixel space, the object loss *ℒ*_*obj*_ was computed for the matched predicted and the ground truth landmark, where *ℒ*_*obj*_ was defined as the sum of the coordinates loss *ℒ*_*coord*_ (3), the confidence loss 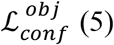, and the classification loss *ℒ*_*cls*_. Since each cell contained at most 2 ground truth landmarks, 3 cases were considered, cells containing 1 landmark, 2 landmarks and no landmark (FIGURE 2).

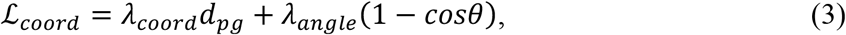

where *ℒ*_*coord*_ denotes the coordinates loss component, *d*_*pg*_ denotes the mean square error (MSE) between the predicted center and the ground truth landmark, *cosθ* denotes the cosine similarity between the predicted and the ground truth vectors, computed by the dot product of their normalized vectors. *λ*_*coord*_ was set to 30.0 and *λ*_*angle*_ was set to 1.0.

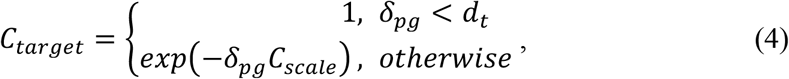

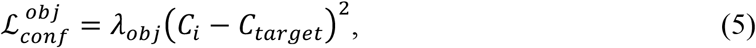

where *C*_*target*_ indicates the target confidence score when ground truth landmark presents, *δ*_*pg*_ was defined as the Euclidean distance in pixel space between the predicted and ground truth landmark. The distance threshold *d*_*t*_ was set to 0.07 and *C*_*scale*_ was set to 2.4. *C*_*target*_ = 1 was only assigned when *δ*_*pg*_ < *d*_*t*_, otherwise the value of *C*_*target*_ varied with *δ*_*pg*_. Such design enabled the correlation behavior between confidence and precision at inference. 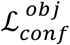 denotes the confidence loss component for cells containing ground truth landmark and *C*_*i*_ corresponds to the predicted confidence score, with *λ*_*obj*_ = 5.0.

As we noticed the difficulty of distinguishing the apex and basal point class because they were located close to each other, focal loss (28) was integrated as *ℒ*_*cls*_, with the parameter *γ* = 2.0 and *α* = (0.4, 0.4, 0.2), respectively for apex, basal point and round window center class, with the *λ*_*cls*_ = 2.0.

The total loss 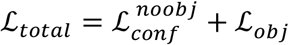, which accumulates the losses of all grid cells in the input image integrating each specific case, providing the objective function for the network training.

### Custom Data Generator

For data preparation, different operations were implemented. To address the issue of strong metal artifact presented by the frame attached on the 3D loaded CT volume, intensity thresholding was applied, where the threshold was determined adaptively by calculating the mean and standard deviation of the voxel intensities in the loaded volume. The voxel values exceeding this threshold were replaced with 0. The entire volume voxel values were then normalized to a range between 0 and 1. Due to the scarcity of manually annotated 3D CT images, and to prevent the potential overfitting (29) issue, a custom 2D data generator was implemented. This generator can produce a large, adjustable amount of training and validation data for the network, containing both 2D cochlea images and background-only images, along with their label matrix. To compensate for the lack of 3D spatial context information resulting from the 2D network design, 3D data augmentation operations were also incorporated into the generator.

For cochlea images, a 3D rigid rotation around the CT volume center was firstly applied: ±6° rotation about x-axis and y-axis, and a wider angular range rotation about the z-axis (within the axial plane) with higher sampling probability within ±30° range. Subsequently, the central axial 2D slice contains the ground truth landmark were extracted from the rotated 3D volume. A translation operation was additionally integrated. For non-rotated volumes, a 2D translation was applied on the extracted axial 2D slice. When involving 3D rotation, a 3D rigid transformation was firstly applied to the CT volume by randomly translating its rotation center within a defined translation range, then the central axial slice was extracted from the transformed volume while ensuring the presence of the landmarks on the extracted 2D slice. The same 3D augmentations were applied to background-only images. However, owing to the substantial imbalance between images containing landmarks and background, the amount of augmented background images was correspondingly reduced.

According to the different dimension of the input CT volume (covering full head or temporal bone region), the extracted 2D slices were cropped or padded to 608 × 608, while preserving at least the full temporal bone region. Furthermore, 2D augmentation operations including Gaussian blur and random sharpness were applied.

For the network’s learning, the ground truth labels for training must align with the augmented images, meaning the labels must correspond to the transformed positions of the target landmarks in the augmented images. To achieve this, the corresponding geometric transformations were also applied to the original ground truth label matrix.

The label matrix was structured the same as the network’s output format, with the shape of 13 × 13 × 2 × 9, which contains coordinates information (x,y,w,h), confidence score, and the class label of 4 classes (one-hot encoding). Thus, this custom data generator was able to flexibly produce augmented 2D images with their corresponding ground truth label matrix, for all 3 landmark classes (apex, basal point and round window center) and a background class.

### Training Configuration

The complete framework was implemented using PyTorch (30) and trained from scratch on 4 NVIDIA Tesla V100 GPUs with 32GB RAM. All of the 4 GPUs were employed to accelerate the training process by integrating Distributed Data Parallel (DDP) via NCCL. To reduce memory consumption, automatic mixed precision (AMP) with FP16 precision was integrated. The training process was configured with 100 epochs, a batch size of 16, and an AdamW (31) optimizer with weight decay set to 0.005. The initial learning rate was set to 0.0001 and the cosine annealing scheduler was enabled. To compare performances of different models, a total of 4 independent models were trained on 4 different training sets, each training set comprises 4, 8, 12 and 16 CT volumes respectively, and the corresponding validation sets is 1,2,3 and 4 CT volumes.

### Localization Inference Pipeline

At inference, the entire 3D CT volume (containing the full head or the temporal bone region) was processed sequentially slice by slice along the axial direction. Each input 2D slice was cropped or padded (without resizing) to 608 × 608 while preserving at least the full temporal bone region. The trained model inferenced on each slice, generating a 13 × 13 × 2 × 9 output tensor. This tensor contains the landmark coordinates information, confidence score, and a class label with one-hot encoding format. Subsequently, the predicted tensors across each axial 2D slice were aggregated together based on their class and the confidence threshold and then clustered in 3D space using DBSCAN (20). The centroid of each cluster was then computed. These centroids were assigned to the left or right side based on their x or y coordinates, which correspond to the anatomical position of the left and right cochleae in the 3D CT scan. This final predicted output label was then compared with the ground truth label, and the localization error was measured using 3D Euclidean distance, in mm. The number of predicted output tensors can vary with the confidence threshold setting, which can potentially influence localization precision.

Thus, in a single forward pass, the proposed network automatically and simultaneously classifies and localizes a total of 6 cochlea landmarks in the input 3D CT image, for each cochlea (left/right cochleae): apex, basal point and round window center.

## RESULTS

To investigate the localization performance, 16 CT volumes from the test set were used. All the CT scans had isotropic image spacing of 0.3 mm. The evaluation consisted of both quantitative and qualitative analyses. Metrics employed include the 3D Euclidean distance and the successful detection rate (SDR@3.0mm), the latter defined as the percentage of detected landmarks with the localization errors bellow 3.0 mm. The localization error was calculated by measuring the 3D Euclidean distance in mm, between the predicted landmark position and its ground truth position in 3D space. The missed landmark localizations were considered as undetected when calculating the SDR@3.0mm. Furthermore, the inference speed was evaluated based on the duration (ms) of single 2D slice inference and the latency (s) of end-to-end entire 3D volume inference, including post-processing. All experiments were performed on an Apple M1 MacBook Pro, 16 GB memory, with Metal Performance Shaders (MPS) enabled for GPU acceleration.

### Few-Volume Learning Analysis

The few-volume learning characteristic of the MEDICO-YOLO was first evaluated by comparing 4 models trained on 4 different training set sizes (respectively 4, 8, 12, 16 CT volumes). The evaluation was contacted on the entire test set of 16 volumes, in total of 96 landmarks, the confidence threshold was set to 0.7 for all the inferences. FIGURE 3 and TABLE 2 demonstrate that the models trained with larger training set size achieve a higher SDR@3.0mm, and reduced dispersion and outliers. Especially, the model achieved high localization accuracy when trained only on 8 CT volumes, with a mean localization error of 1.30 mm (approximately 4 voxels). While training on 16 CT volumes, the model achieved peak localization performance among the evaluated training set sizes, with SDR@3.0mm of 97.9 %, and an overall mean localization error of 0.71 mm (approximately 2.4 voxels). Specifically, 0.85 mm for the apex, 0.67 mm for the basal point, and 0.60 mm for the round window center. This additionally indicates the localization performance varied across different anatomical landmarks, nevertheless, sub-millimeter precision was achieved for all landmarks. These findings demonstrate that the MEDICO-YOLO framework achieved promising localization performance from few annotated ground truth 3D CT scans. This few-volume learning capacity may benefit from the synergistic design of the entire framework, particularly the custom data generator with its data augmentation.

**TABLE 2.** Comparison of localization performance across different training set sizes. Apex: helicotrema; Basal: center of the basal turn of the cochlea; RW: round window center; SDR@3.0mm(%): percentage of successfully detected landmarks within error threshold of 3 mm.

| **Model (Training Set Size)** | **Overall Mean Distance (mm)** | **Apex (mm)** | **Basal (mm)** | **RW (mm)** | **SDR@3.0 mm(%)** |
|---|---|---|---|---|---|
| 4 volumes | 2.28±1.62 | 2.64 | 2.48 | 1.79 | 55.21 |
| 8 volumes | 1.30±0.86 | 1.45 | 1.40 | 1.05 | 70.83 |
| 12 volumes | 1.04±0.50 | 1.16 | 1.07 | 0.87 | 79.18 |
| **16 volumes** | **0.71±0.33** | **0.85** | **0.67** | **0.60** | **97.92** |

**FIGURE 3.**
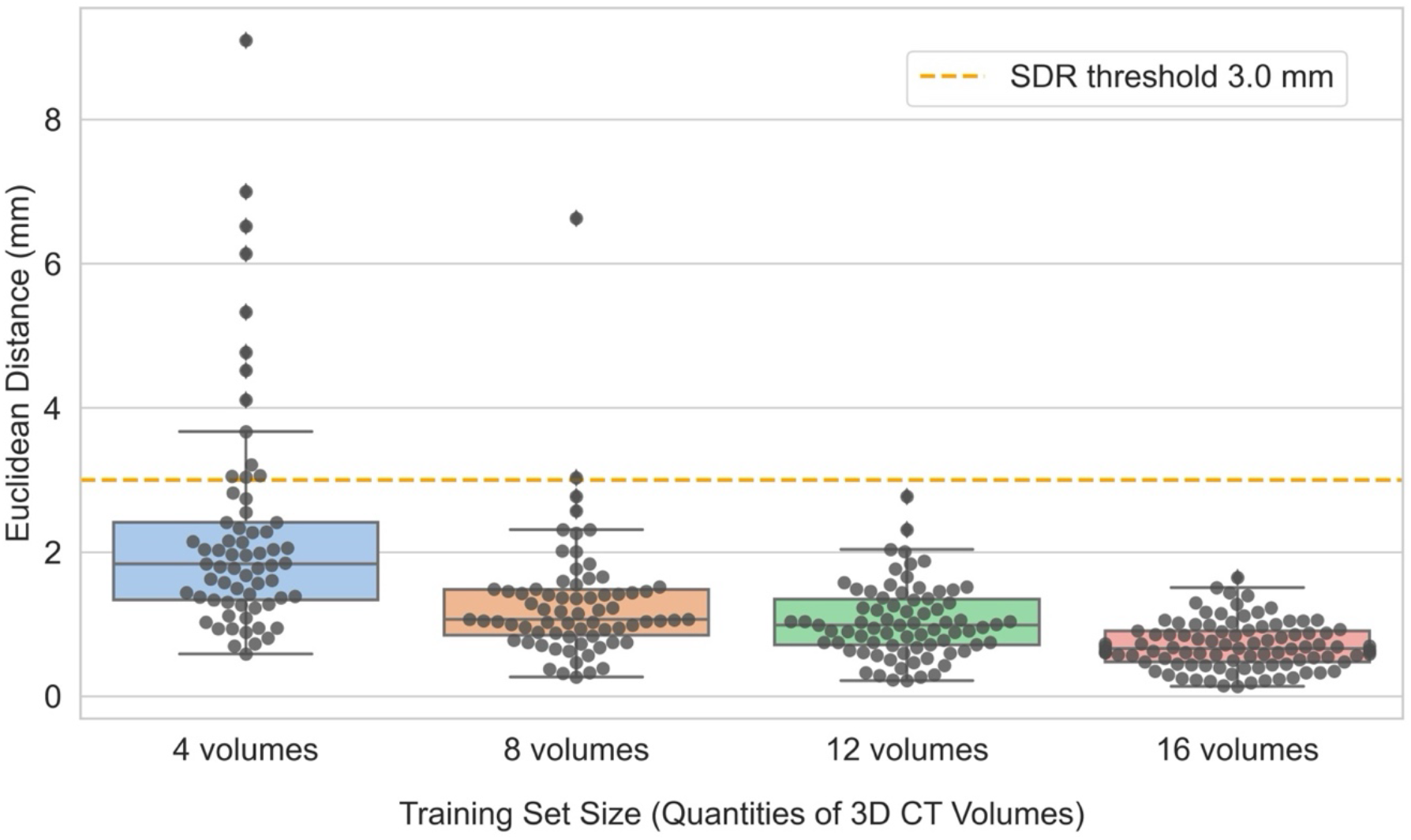
Localization performance comparison (mm) across varying training set sizes. The x-axis indicates model versions trained on different training set. The y-axis indicates the 3D Euclidean distance (mm) between predicted and ground truth landmark coordinates. Each dot represents the localization error for a single landmark predication. Missed landmark localizations were excluded.

### Localization Inference (Speed & Precision) Across Different Confidence Thresholds

To further investigate the impact of confidence thresholding, the model trained on 16 volumes was assessed on the test set, under different confidence thresholds (TABLE 3). Mean distance, SDR@3.0mm and inference speed per slice (2D) and per volume (3D), were measured over all 16 test volumes. We report to 3 decimals to highlight the precision difference under varied confidence thresholds. Results demonstrate the precision improved slightly as confidence threshold increases, reaching its peak at confidence threshold 0.9, where localization error attained 0.678 mm. SDR@3.0mm decreases at higher confidence threshold, but all beyond 90%. This precision confidence threshold behavior indicates a trade-off exists between localization error and SDR@3.0 mm, where larger confidence threshold results in more precise detection but a lower SDR@3.0 mm.

**TABLE 3.**
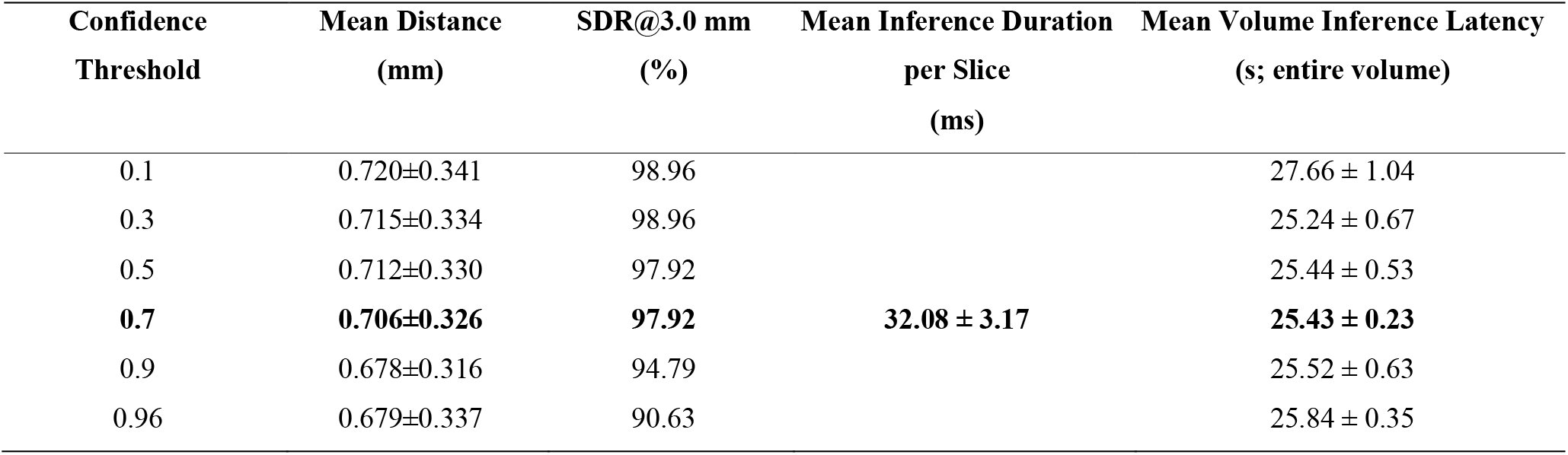
Localization performance across different confidence thresholds. Inference per Slice: forward inference duration on single slice; End-to-end volume inference latency: total inference duration on a 3D CT volume (incl. image preprocessing, network inference, and post-processing).

| Confidence Threshold | Mean Distance<br>(mm) | SDR@3.0 mm<br>(%) | Mean Inference Duration per Slice<br>(ms) | Mean Volume Inference Latency<br>(s; entire volume) |
|---|---|---|---|---|
| 0.1 | 0.720±0.341 | 98.96 | **32.08 ± 3.17** | 27.66 ± 1.04 |
| 0.3 | 0.715±0.334 | 98.96 |  | 25.24 ± 0.67 |
| 0.5 | 0.712±0.330 | 97.92 |  | 25.44 ± 0.53 |
| **0.7** | **0.706±0.326** | **97.92** |  | **25.43 ± 0.23** |
| 0.9 | 0.678±0.316 | 94.79 |  | 25.52 ± 0.63 |
| 0.96 | 0.679±0.337 | 90.63 |  | 25.84 ± 0.35 |

As confidence thresholding doesn’t impact the network’s each forward inference computation, the average inference duration per 2D slice remains consistent at approximately 32 ms across different confidence thresholds (Apple M1 MacBook Pro). In parallel, we measured the model’s end-to-end inference latency per volume, defined as the duration of the complete inference process for the input 3D CT volume. This includes preprocessing the loaded CT, network forward inference of all the 2D slices, and the post-processing allows to output the definitive coordinates of 6 landmarks in the 3D CT volume (both left and right cochleae). Since end-to-end volume inference latency (3D) varies with the volume depth, 10 representative CT volumes from test set with the same depth of 200 slices were employed. TABLE 3 shows that different confidence thresholds lead to subtle variations in latency, this is because lower confidence threshold outputs more detected coordinates, thus slightly increases post-processing computation. When the confidence threshold set at 0.7, the model completed the localization of full-resolution, 200-slice depth 3D CT volumes with a mean latency of 25.4 s (Apple M1 MacBook Pro laptop). Together, the proposed MEDICO-YOLO framework performs real-time inference on each 2D slice and completes localization of the entire 3D CT volume in under half a minute while attaining high precision.

### Qualitative Localization Visualization

To qualitatively evaluate the localization performance, the output of the model trained on 16 volumes was compared with human expert’s annotation (FIGURE 4) on a representative 3D CT image from the test set. We can visually observe that the model-predicted landmarks closely overlap (within around 3 voxels) with the ground truth landmarks.

**FIGURE 4.**
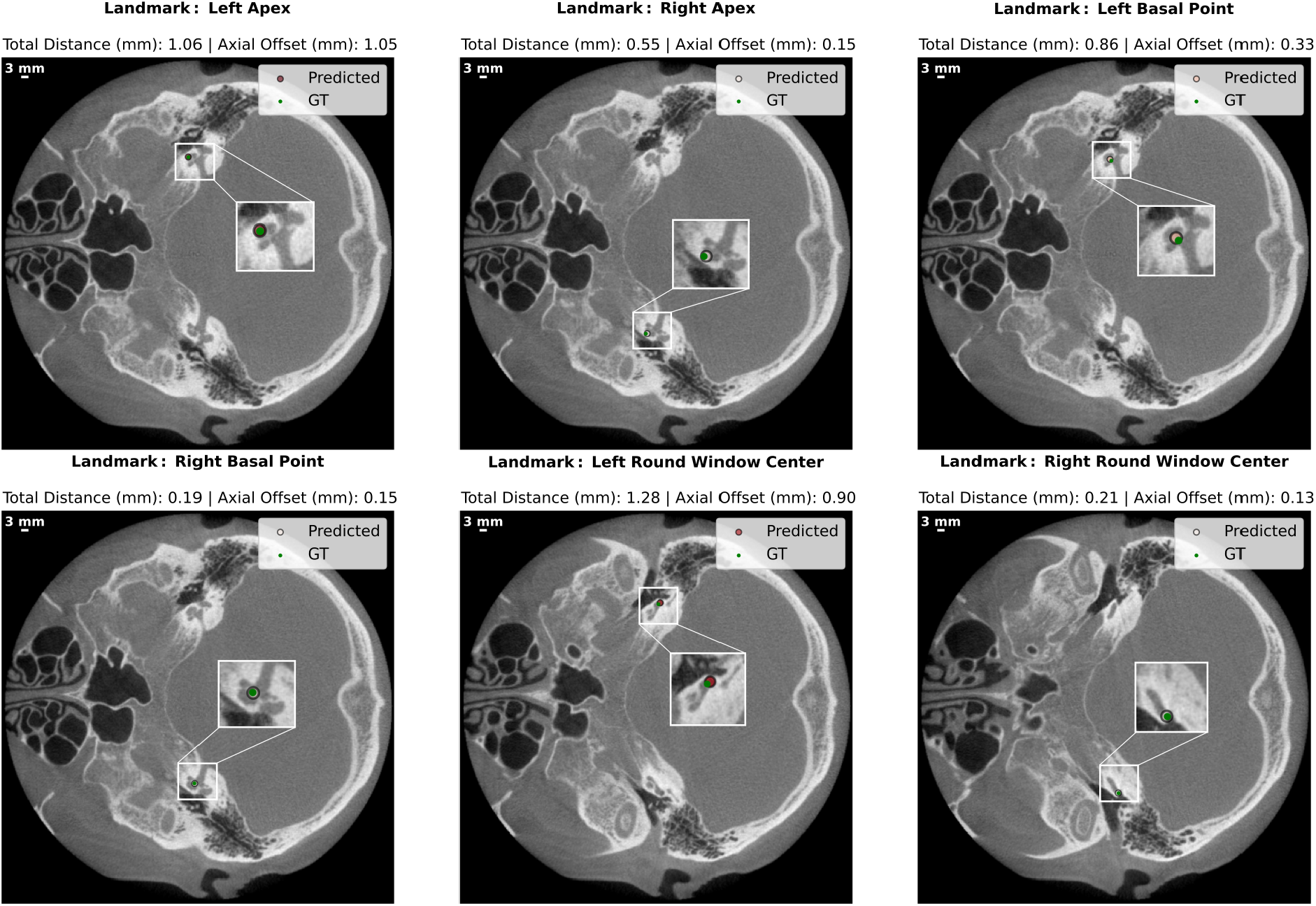
Visual comparison between ground truth landmarks and predicted landmarks of 16 volumes-trained model (confidence threshold 0.7) on a representative full temporal bone CT scan from test set (volume shape: 536 × 536 × 198) in axial view. For each landmark, total distance represents the Euclidean distance in 3D space, and axial offset represents error along the axial direction, while the color variation of predicted landmarks illustrates the magnitude of axial offset. The mean 3D localization error for all the landmarks on the presented CT scan is 0.69 ± 0.41mm.

### Localization Precision Comparison with Human Experts

In particular, we also quantitatively compared the proposed framework with the human experts’ annotations in terms of the precision (FIGURE 5). 3 human experts manually labeled 10 cochleae on a different dataset than the model’s test set (but the same acquisition source). One expert’s annotation served both as the ground truth for the model and as the reference for comparing with the other 2 human experts’ annotations. The model’s localization precision was evaluated on the same 16 test volumes used in FIGURE 3 (in total of 32 cochleae). The missed landmark predictions from the model were excluded, which is consistent with SDR@3.0mm of 97.9 %. FIGURE 5 illustrates a minor inter-expert variability, while the model achieved a level of localization precision comparable to that of human experts in this particular task (localizing 3 landmarks of both left and right cochleae). To further investigate, a Mann-Whitney U test was performed, owing to the different datasets annotated by the human experts and the model. The output suggests that for overall classes, the model’s localization errors are still significantly different from those of the human experts (*p* = 0.024). However, specifically for localizing the round window center, the model’s localization errors are not significantly different from those of the human experts (*p* = 0.707). However, this statistical observation can be limited due to the scarcity of human expert annotation resources.

**FIGURE 5.**
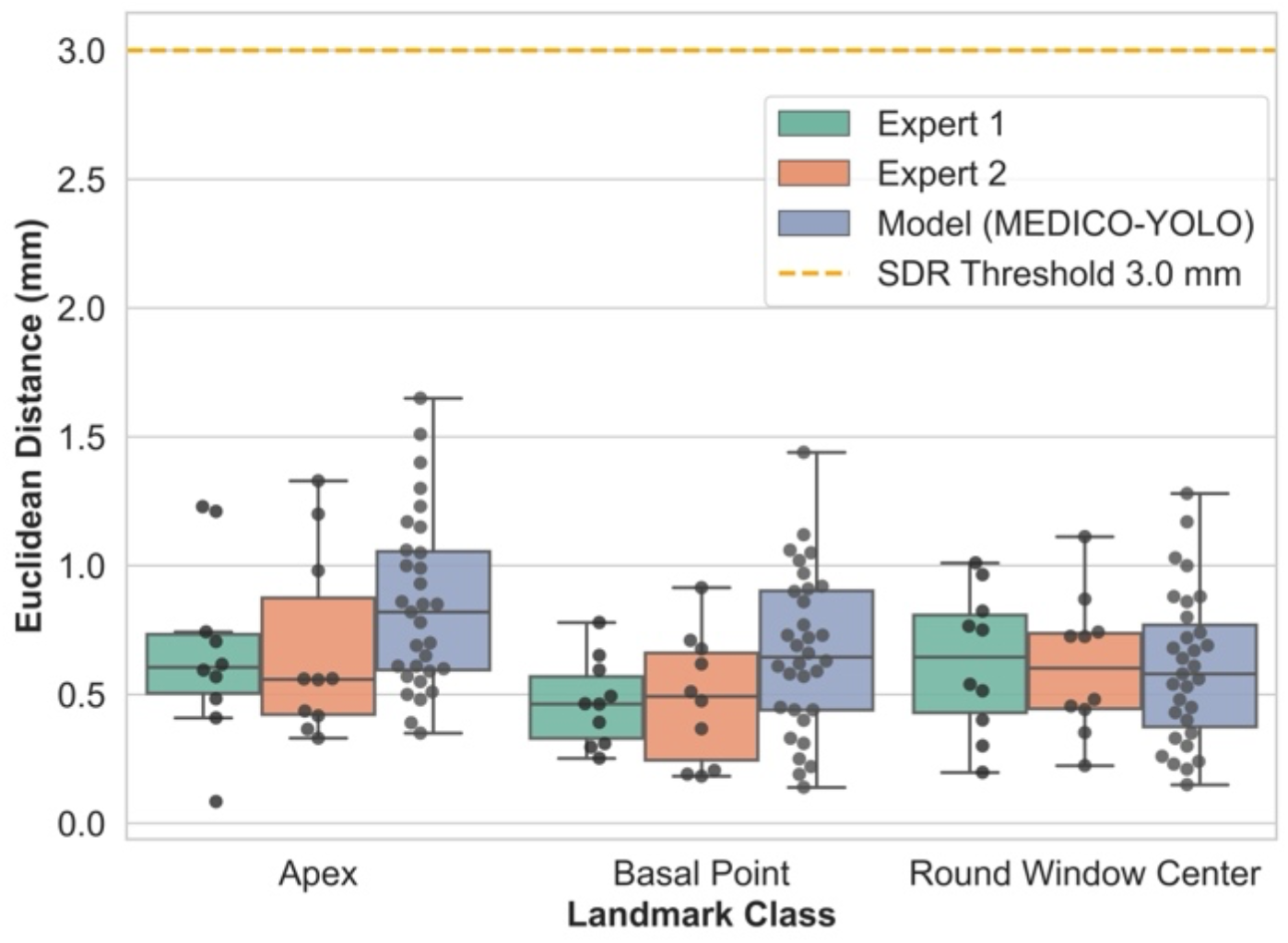
Localization error comparison between the model and the human experts. The x-axis indicates different landmark classes. The y-axis indicates the Euclidean distance (mm) in 3D space between predicted landmark position and its true position. Model employed is the 16 volumes-trained version with confidence threshold set at 0.7. Each dot represents the localization error for a single landmark annotation.

## DISCUSSION

### Overall Comparison with Related Works

To provide a general view of the field, we compared the proposed framework with the other relevant studies in terms of overall methodology (TABLE 4). Due to the differences in datasets and methods, this comparison was performed using the results reported in the respective literature, only the localization part was compared. Each method presents distinct advantages in its specific clinical scenario. Wang et al. (14) achieved CNN based landmark localization with small-scale training dataset, each trained model demonstrated robust localization precision. Stebani et al. (13) presented the precise localization of structures including the oval window. Wang et al. (12) introduced an approach that only a single labeled CT image was employed for the training. Margeta et al. (11) developed an approach that capable of localizing landmarks on both pre- and post-operative CT scans, thereby offering a more general usage scenario. Meanwhile, in this specific task of localizing the landmarks of both left and right cochleae on pre-operative 3D CT scans, the advantage of our approach is that it achieved a balance between the few-volume learning, a fully automated localization pipeline, the rapid inference speed, and high localization precision.

**TABLE 4.** Overall Comparison with Other Related Works in Temporal Bone Landmarks Localization. NR: not reported; ROI: region of interest; SOI: structure of interest; Pre-operative CT scan: scans without electrode implanted.

| Methods | Mean Localization Error (Test set) | Training Set Size | Inference Latency (Device) | Manual Initialization/Sub-volume/ROI/SOI | Target Landmarks | Image Type |
|---|---|---|---|---|---|---|
| Wang et al. [14] | 0.9 mm<br>(3 voxels) | 1 to 11 | NR | Yes<br>(40 x 40 x 40 voxels, expandable ROI with increased latency) | Apex, basal point, round window | Pre-operative CT scan |
| Stebani et al. [13] | 5.2 voxels<br>(isotropic voxel edge length 99µm) | 38 | 50s, TTA strategy<br>(NR) | Yes<br>(128 x 128 x 128 voxels) | Helicotrema, oval window, round window | Pre-operative CT scan |
| Wang et al. [12] | Around 0.6 mm<br>(2 to 3 voxels) | 1 | 30s<br>(NVIDIA 1080 Ti GPU) | Yes<br>(SOI + registration) | Cochlea Center, top, window | Pre-operative CT scan |
| Margeta et al. [11] | 0.71/0.75/1.30 mm<br>(per landmark, isotropic 0.3 mm spacing) | 279 | 5.9s<br>(Azure Cloud VMs, Standard DS3 V2) | Yes<br>(10mm x 10mm x 10 mm) | Apex, center, round window | Pre- and post-operative CT scan |
| **Proposed method** | **0.71 mm<br>(2.4 voxels)** | **4 to 16** | **25s<br>(Apple MacBook Pro M1)** | **No<br>(608 x 608 x 200 voxels)** | **Apex, basal point, round window center** | **Pre-operative CT scan** |

Overall, achieving such encouraging performance underscores the strength of the framework’s synergistic and task-oriented design, which encompasses the network architecture, loss function, data generation, training configuration, and inference pipeline. Each component is tailored to the others, collectively forming an end-to-end localization framework.

### Limitations and Future Work

This work has several limitations. First, since the data generator was designed for generating axial slices from the 3D CT scans, the model can only process the input 3D CT in the axial direction. Second, only the anatomical landmarks of the cochlea structure were localized, whereas additional structures are also interesting to involve for more comprehensive surgical plannings. Another limitation is that this study integrated exclusively pre-operative CT scan data, which restricts its utility in post-operative analysis. In future work, we aim to directly address these limitations by: (1) expanding the data generator to include further sagittal and coronal view slices, (2) extending the framework to localize landmarks of additional structures (e.g., ossicles, facial nerve) without largely modifying the design principle of the MEDICO-YOLO framework, and (3) involving post-operative CT scans through metal artifact reduction or registration techniques. This work can also be extended by integrating multi-modal imaging data.

## CONCLUSION

In conclusion, both quantitative and qualitative experiments demonstrated that the proposed MEDICO-YOLO framework can localize multiple cochlea landmarks in 3D pre-operative CT scans fully automatically and with reliable localization performance, thereby supporting its practical integration into pre-operative surgical planning for cochlear implant surgery. A broader clinical application can also be envisaged with the planned expansion in future work.

## AUTHOR CONTRIBUTIONS

**Y.W:** Writing — original draft, Writing — review & editing, Data curation, Investigation, Methodology, Formal analysis, Software, Visualization, Validation, Conceptualization. **A.K:** Conceptualization, Supervision, Writing — review & editing. **T.L:** Conceptualization, Supervision, Writing — review & editing, Funding acquisition, Resources. **S.J:** Supervision, Writing — review & editing, Resources, Conceptualization, Validation, Funding acquisition, Project Administration.

## ETHICAL APPROVAL AND CONSENT TO PARTICIPATE

The ethical approval regarding anonymized data was confirmed by Medizinische Hochschule Hannover with the institutional review board’s generic ethics decision No. 1897-2013 from 2013-07-09 regarding publications with patient data, and by University of Rostock with ethics vote No. A 2016-0083.

## ACKNOWLEDGMENTS

The authors would like to thank the support from Comm4CHILD project and Medizinische Hochschule Hannover. We also would like to thank Dr. Felix Repp for the discussion of transformation matrices within the CT and the manual annotation.

## DECLARATION OF AI ASSISTED TOOLS

DeepL tool was used only for language refinement of the text. The author reviewed, refined and validated all the text. The author takes full responsibility for all the content of this work.

## DATA AVAILABILITY STATEMENT

Part of the generated or analyzed data in this study can be shared by the authors on reasonable requests.

